# A Cross-Sectional Examination of Conflict-of-Interest Disclosures of Physician-Authors Publishing in High-Impact US Medical Journals

**DOI:** 10.1101/2021.09.12.21263468

**Authors:** James H. Baraldi, Steven A. Picozzo, Jacob C. Arnold, Kathryn Volarich, Michael R. Gionfriddo, Brian J. Piper

## Abstract

**Objective:** To assess the accuracy of self-reported financial conflict-of-interest (COI) disclosures in the *New England Journal of Medicine* (*NEJM*) and *Journal of the American Medical Association* (*JAMA*) within the requisite disclosure period prior to article submission.

**Design:** Cross-sectional investigation.

**Data Sources:** Original clinical-trial research articles published in *NEJM* (*n* = 206) or *JAMA* (*n* = 188) from January 1 to December 31, 2017; self-reported COI disclosure forms submitted to *NEJM* or *JAMA* with the authors’ published articles; Open Payments website (from database inception; latest search: August 2019).

**Main outcome measures:** Financial data reported to Open Payments from 2014 to 2016 (time period that included all subjects’ requisite disclosure windows) were compared to self-reported disclosure forms submitted to the journals. Payments were defined as those not associated with a research study or formal research funding. Payment types were categorized as “disclosed,” “undisclosed,” “indeterminate,” or “unrelated.”

**Results:** Thirty-one articles from *NEJM* and 31 articles from *JAMA* met inclusion criteria. The physician-authors (*n* = 118) received a combined total of $7.48 million. Of the 106 authors (89.8%) who received payments, 86 (81.1%) received undisclosed payments. The top 23 most highly compensated received $6.32 million, of which $3.00 million (47.6%) was undisclosed. Disclosure rates were the equivalent between the top 23 and the entire sample.

**Conclusions:** High payment amounts, as well as high proportions of undisclosed financial compensation, regardless of amount received, comprised potential COIs for two influential US medical journals. Further research is needed to explain why such high proportions of general payments were undisclosed and whether journals that rely on self-reported COI disclosure need to reconsider their policies.

## BACKGROUND

Financial conflicts of interest (COIs) are a perennial problem for medical research and practice.^1,2^ Physician researchers who receive industry payments are more likely to demonstrate results favorable to the companies funding them;^3,4^ are more likely to prescribe drugs and use of medical devices produced by these companies, from statins^5^ to opioids^6^ to endoscopic^7^ and orthopedic devices;^8^ and they may unduly influence other physicians by contributing to research that others use to guide their own clinical practice.^9-16^ Industry payments to physicians therefore may bias healthcare providers’ delivery of evidence-based medicine and interfere with their responsibilities to their patients.

In order to increase the transparency of the financial relationships between physicians and pharmaceutical and medical device manufacturers, the US government passed the *Physician Payment Sunshine Act* as part of the *Patient Protection and Affordable Care Act* in 2010.^17^ This law required manufacturers reimbursed by Medicare, Medicaid, or the Children’s Health Insurance Program to submit information regarding payments received by physicians to the Centers for Medicare and Medicaid Services (CMS). CMS shares these payment data with the public on an annual basis through the Open Payments website,^18^ which was introduced in 2014 with data starting in August of 2013. The International Committee of Medical Journal Editors (ICMJE) has produced its own COI form to help medical journals maintain COI disclosure standards for physician-authors seeking to publish articles in peer-reviewed medical journals.^18^ Many journals have adopted the use of this form, requiring authors submitting manuscripts to them to disclose payments received from manufacturers of products related to the article content in the 36 months prior to submission.^19^ These author disclosures can be verified by viewing the physician-author’s record in the Open Payments database.^18^

Despite these attempts to address COI disclosures, COI disclosure opacity has persisted across a diversity of specialties,^20,21^ forms of compensation,^22^ and investigational products of clinical trials.^23-26^ Previous studies of inaccurate, or “discordant,” COI disclosures have emphasized financial COIs,^27^ COIs differing significantly by specialty,^28^ and inaccurate COI disclosures appearing in high-impact journals, such as the *New England Journal of Medicine* (*NEJM*).^25^ In 2017 the *Journal of the American Medical Association* (*JAMA*) published an issue dedicated to the subject of COI disclosures to highlight the multifaceted nature of the problem.^29^

*NEJM* and *JAMA* are the peer-reviewed general medical journals published in the United States with the highest and second-highest impact factors, respectively. Both journals publish with similar frequency (weekly for the former and 48 times per year for the latter), emphasize publication of original research as well as reviews, are popular for physician-authors, and publish articles that receive wide coverage both within the scientific community and in the popular news media. The impact and reach of these two journals have substantial potential to shape future research and patient care. To date, there has been no comprehensive study of COI disclosures in these two journals. This study examined COI disclosures among physician-authors who published articles in either (or both) of these journals in 2017, the first year for which complete data exist for the earliest possible disclosure period following the inception of Open Payments. Identifying patterns of disclosure transparency at the beginning of the existence of Open Payments revealed the extent to which physician-authors publishing in *NEJM* and *JAMA* follow COI disclosure policies.

## METHODS

### Inclusion Criteria

Original research articles (*n* = 394) detailing the results of randomized clinical trials and published in *NEJM* (*n* = 206, 52.3%) and *JAMA* (*n* = 188, 47.7%) from January 1, 2017, to December 31, 2017, were examined. The first and last author of each article were identified. Articles were excluded from further examination if either the first or last author did not have an M.D. or D.O. degree and if either author did not have a record in Open Payments. These authors had to be physicians with Open Payments records. This is due to this study’s objective of exploring COI disclosures of physician-authors by comparing Open Payments records, which consisted of disclosures made by the companies making the payments, with self-disclosures made by the authors to the journals. These self-disclosures consisted of the authors’ identifications of the companies that paid them, not of the amounts, which the companies themselves provided to Open Payments.

### Data Collection

Data reported to Open Payments from 2014 to 2016 were compiled and compared to self-reported disclosure forms that had been submitted to *NEJM* or *JAMA* with the authors’ published articles. Data collected about the authors included sex, specialty, journal(s) of publication, and yearly payment information. Open Payments defines “payments” as either general “payments that are not associated with a research study” or research payments that “are associated with a research study.”^30^ This study focused solely on general payments, which included compensation for promotional speaking, consulting, travel and lodging, food and beverage, honoraria, and current or prospective ownership or investment interest.

The articles were examined to determine their areas of investigation (e.g., cardiovascular disease, diabetes, cancer). This occurred with reference to the title, key words, abstracts, and content of each article. Three co-authors of this study collected the data, resolving by discussion any disagreement in interpretation of article topics for the purpose of disclosure analysis. Each article’s area of investigation was compared against the product portfolios and research pipelines of the companies that paid the physician-authors, according to their Open Payments data. Payments from companies disclosed by the author were labeled as “disclosed” for this study. Payments from companies not listed on the respective authors’ disclosure forms were investigated further and categorized as “undisclosed,” “indeterminate,” or “unrelated.” Occasionally, a company that did not match the disclosures on the author’s form was later determined to have made a disclosed payment. For example, a company with multiple names was determined to have made a disclosed payment under only one name, prompting a review that revealed the fact that the company’s payment had been disclosed, just under a name that was not immediately recognizable on the disclosure form. Every payment from a given company was analyzed by researching that company, thus putting all payments from that company under one umbrella for the purpose of COI disclosure. For example, if a company produced a drug related to the content of an author’s research article, then every general payment that that company made to the author, regardless of the nature of the payment, was construed as a COI. Payments were considered for both the parent and subsidiary companies.

The criteria in Table 1 were followed to categorize payments as “disclosed,” “undisclosed,” “indeterminate,” or “unrelated.” These definitions were adapted from the ICMJE disclosure form used by both journals, which states:

**TABLE 1.** Disclosure category descriptions and examples, per ICMJE guidelines. *An individual physician-author can dispute a payment; therefore, this amount would not have to be disclosed if the physician-author believes that he/she had not received it.

| **Payment Category** | **Definition** | **Example** |
| --- | --- | --- |
| Disclosed | A payment was considered disclosed if the author disclosed a payment from a company that matched the data from Open Payments. | A physician-author was doing research on cancer and reported a payment from a company that has several chemotherapeutic patents in its portfolio. |
| Undisclosed | A payment was considered undisclosed if:<br>1) the author received a payment during the relevant disclosure period that did not match any disclosures provided to the journal, AND<br>2) the company offers, or offered at the time of the payment, a product that could broadly be considered related to the area of inquiry. | A physician-author was doing research on cardiovascular disease, received a payment from a company that produces anti-hypertensive medication and that was not listed on the disclosure form, and did not report the payment from that company on the author disclosure form. |
| Indeterminate | A payment was considered indeterminate if:<br>1) the author received a payment during the relevant disclosure period that did not match any disclosures provided to the journal, BUT<br>2) the company was a subsidiary or parent company of a company listed on the disclosure, AND/OR<br>3) it could not be determined whether that company offers, or offered at the time of the payment, a product that could broadly be considered related to the area of inquiry, AND/OR<br>4) the payment has been disputed.* | (A) The physician-author was doing research on a new surgical product, reported a payment from Johnson & Johnson, and Open Payments listed a payment from Ethicon, a subsidiary of Johnson & Johnson.<br>(B) The physician-author was doing type I diabetes research, and a company has type II diabetes products. |
| Unrelated | A payment was considered unrelated if:<br>1) it was not disclosed, AND<br>2) the company from which the payment originated does not offer a product that could broadly be considered related to the area of inquiry. | An author in an orthopedic research study is funded by a company that provides heart monitoring technology exclusively. |

> [Authors] should disclose interactions with ANY entity that could be considered broadly relevant to the work. For example, if your article is about testing an epidermal growth factor receptor (EGFR) antagonist in lung cancer, you should report all associations with entities pursuing diagnostic or therapeutic strategies in cancer in general, not just in the area of EGFR or lung cancer.^31^

ICMJE requires disclosures for 36 months prior to submission. Therefore, this study focused on all payments within 36 months of the submission dates. A copy of the ICMJE form reviewed for this study is available in the Supplementary Materials.^31^

Payment data in the study window, including company name, amount, and purpose, were manually extracted from the Open Payments database into a spreadsheet accessible to all authors of this study. Payments were then categorized based on the ICMJE guidelines (Table 1) by three co-authors in their first two years of medical school and disagreements were resolved by discussion among these raters. *NEJM* provided the disclosure forms as attachments to their articles; *JAMA* provided a list of disclosures at the end of each article. The *NEJM* articles investigated in this study stated their submission dates or included this information on the author disclosure forms, which are made available to the public as attachments to each article. For *JAMA* the submission date was approximated by using the date when the article was published. *JAMA*’s official position is that, as of 2016, the median time from article submission to acceptance was eighteen days, and the median time from acceptance to first online publication another fourteen days, roughly totaling one month.^32^ Therefore, unknown submission dates were estimated as 30 days prior to respective publication dates; this caveat is important for data interpretation. It was assumed that a COI encountered within these thirty days would be unlikely to influence the manuscript, presumably already written in its nearly final form.

Data were collected for payments from 2014 to 2016. Data in Open Payments are periodically updated. Our data were last updated in August of 2019. See the Supplementary Materials for the full data. Our framework conceptualized a broadly construed COI, rather than impact on research per se.

This study was submitted to the Institutional Review Board of the Geisinger and deemed exempt.

### Statistics

Analysis focused on payments received within the years 2014, 2015, and 2016, as all of the authors’ respective 36-month disclosure windows overlapped these years. Additional data outside of the authors’ disclosure windows were collected and are reported in the Supplementary Materials.

GraphPad Prism (version 9) was used for statistical analysis and for figure generation. Descriptive statistics were calculated. ROUT analysis (maximum desired false discovery rate Q = 1%) identified outliers. The Wilcoxon rank-sum test assessed the extent of parity between distributions. The flowchart was generated using Lucidchart (Lucid Software Inc., South Jordan, UT, USA). A *p*-value of *p* < 0.05 was considered statistically significant. Mean amounts are presented ± standard deviation.

## RESULTS

A total of 394 original research articles published in *NEJM* (*n* = 206) and *JAMA* (*n* = 188) from January 1, 2017, to December 31, 2017, were examined. Thirty-one articles from *NEJM* and 31 articles from *JAMA* met all criteria for inclusion, with a total of 118 unique authors.

Within their respective 36-month disclosure windows, the 118 authors received $7,476,049.87 in general payments combined. Payments to authors who published in *NEJM* totaled $3,635,791.81 (48.4% of the total) and to *JAMA* totaled $3,876,107.75 (51.6%). The median payment amount for *NEJM* authors was $11,224.53; at Q1 (25^th^ percentile) the amount was $755.67, and at Q3 (75^th^ percentile) was $80,179.56. For *JAMA* authors, the median payment was $2,400.00, with Q1 at $65.20 and Q3 at $30,964.21. Mean payment amounts were $58,641.80 (±$102,337.65) for *NEJM* and $68,001.89 (±$215,813.16) for *JAMA* (Figure 1). Authors in aggregate received comparable total payments as well as similar total payments by category as proportions of total payments by journal (Table 2 and Figure 2).

**TABLE 2.** Payment amounts by category compared between the New England Journal of Medicine (*NEJM)* and the Journal of the American Medical association (*JAMA)*. Amounts shown sum by column to “Total” and by row to “*NEJM* + *JAMA*.” Percentages shown sum per column. One author (#34) published in both journals; this author’s payment amounts ($35,849.69) were counted once in calculating total general payments but twice (once per journal) for between-journal analysis.

| **Payment Category** | ***NEJM*** | ***JAMA*** | ***NEJM + JAMA*** |
| --- | --- | --- | --- |
| Disclosed | \$2,174,199.92<br>(59.8%) | \$1,675,846.29<br>(47.6%) | \$3,850,046.21<br>(51.3%) |
| Undisclosed | \$1,399,156.79<br>(38.5%) | \$2,027,672.77<br>(48.3%) | \$3,426,829.56<br>(45.6%) |
| Indeterminate | \$22,610.21<br>(0.6%) | \$3,863.42<br>(0.1%) | \$26,473.63<br>(0.4%) |
| Unrelated | \$39,824.89<br>(1.1%) | \$168,725.27<br>(4.0%) | \$208,550.16<br>(2.8%) |
| Total | \$3,635,791.81<br>(100%) | \$3,876,107.75<br>(100%) | \$7,511,899.56<br>(100%) |

**FIGURE 1.**
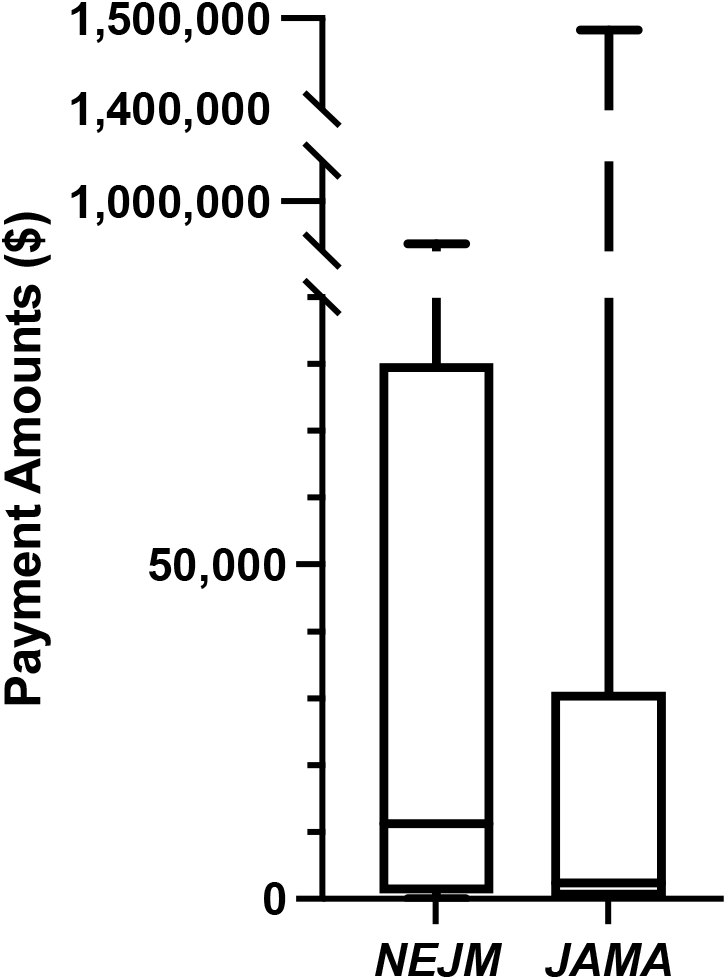
Distribution of total payment amounts compared between *NEJM* and *JAMA. NEJM* authors had a higher median payment amount, but *JAMA* authors had a higher mean. Distribution by COI disclosure rate (analysis not shown) followed a similar pattern.

**FIGURE 2.**
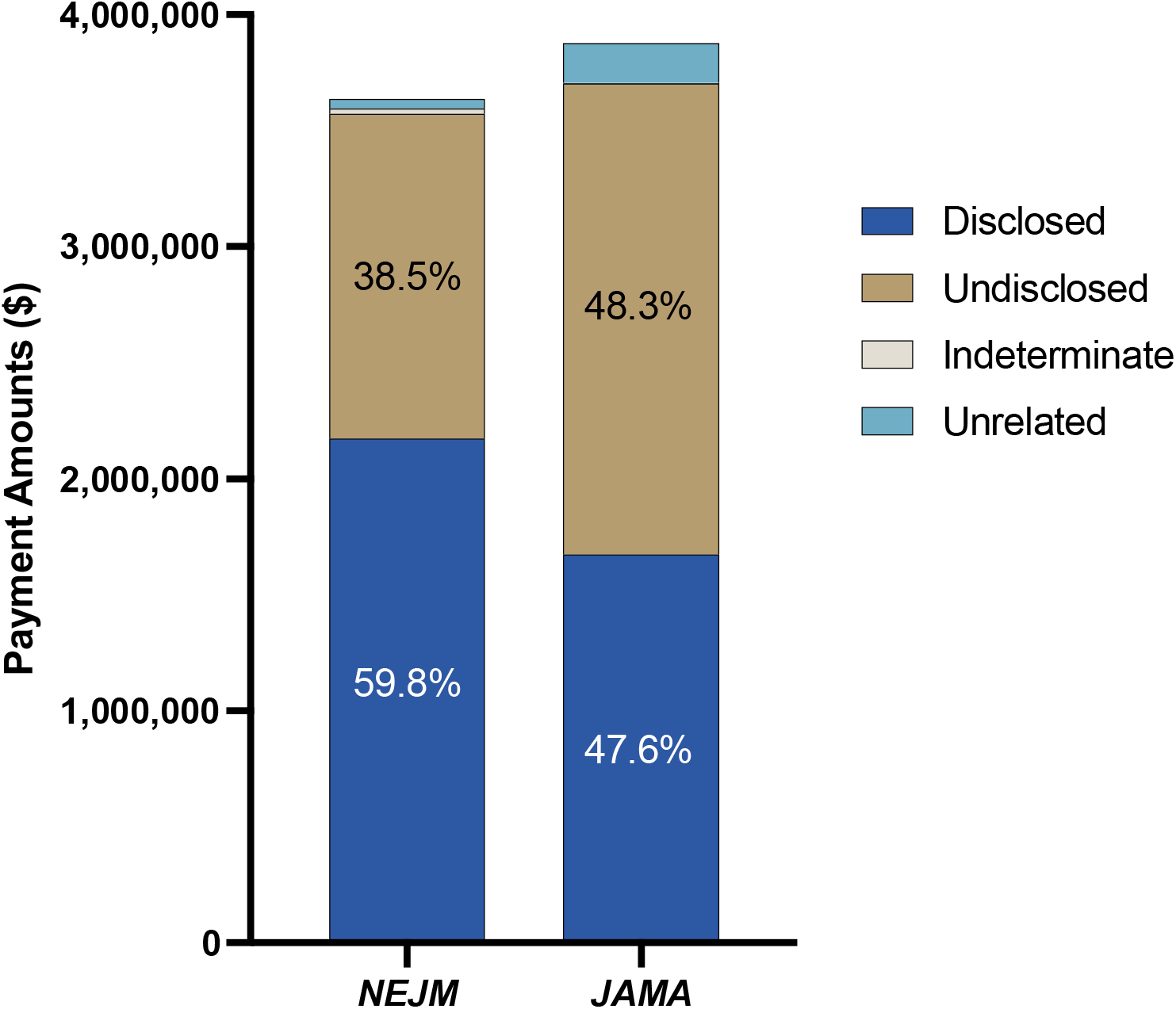
Payment amounts by category compared between *NEJM* and *JAMA*. Percentages shown are of the labeled category with respect to overall payments to those authors publishing in the respective journals. For more detailed information see Table 2.

Of the 118 authors, twelve (10.2%) received no payments. Of the 106 (89.8%) who did, payment amounts ranged from a minimum of $6.36 to a maximum of $1,486,929.34. 86 of these 106 authors (81.1%) received undisclosed payments. Twenty-three outliers were identified, ranging from $93,165.88 to $1,486,929.34, reflecting the payment amounts received by the 23 most highly compensated physician-authors. All 23 had MD degrees; three additionally had PhDs, and two others additionally had MPHs. Sixteen (69.6%) were internal medicine specialists or subspecialists. Fifteen (65.2%) published in *NEJM*, and eight (34.8%) published in *JAMA*. Twelve (52.2%) were first authors, and eleven (47.8%) were last authors. Twenty-one (91.3%) were males, and two (8.70%) were females. The top 23 most highly compensated received $6,316,025.03, of which $3,004,703.54 (47.6%) was undisclosed. The total amount that the top 23 most highly compensated physician-authors received ($6,316,025.03) comprised 84.5% of all compensation received by all 118 physician-authors ($7,476,049.87) (Table 3). The total amount that the *NEJM* outliers received ($2,965,974.61) comprised 81.6% of all compensation received by all 62 *NEJM* authors, and the total amount that the *JAMA* outliers received ($3,350,050.42) comprised 86.4% of all compensation received by all 57 *JAMA* authors. One author published in both journals.

**TABLE 3.**
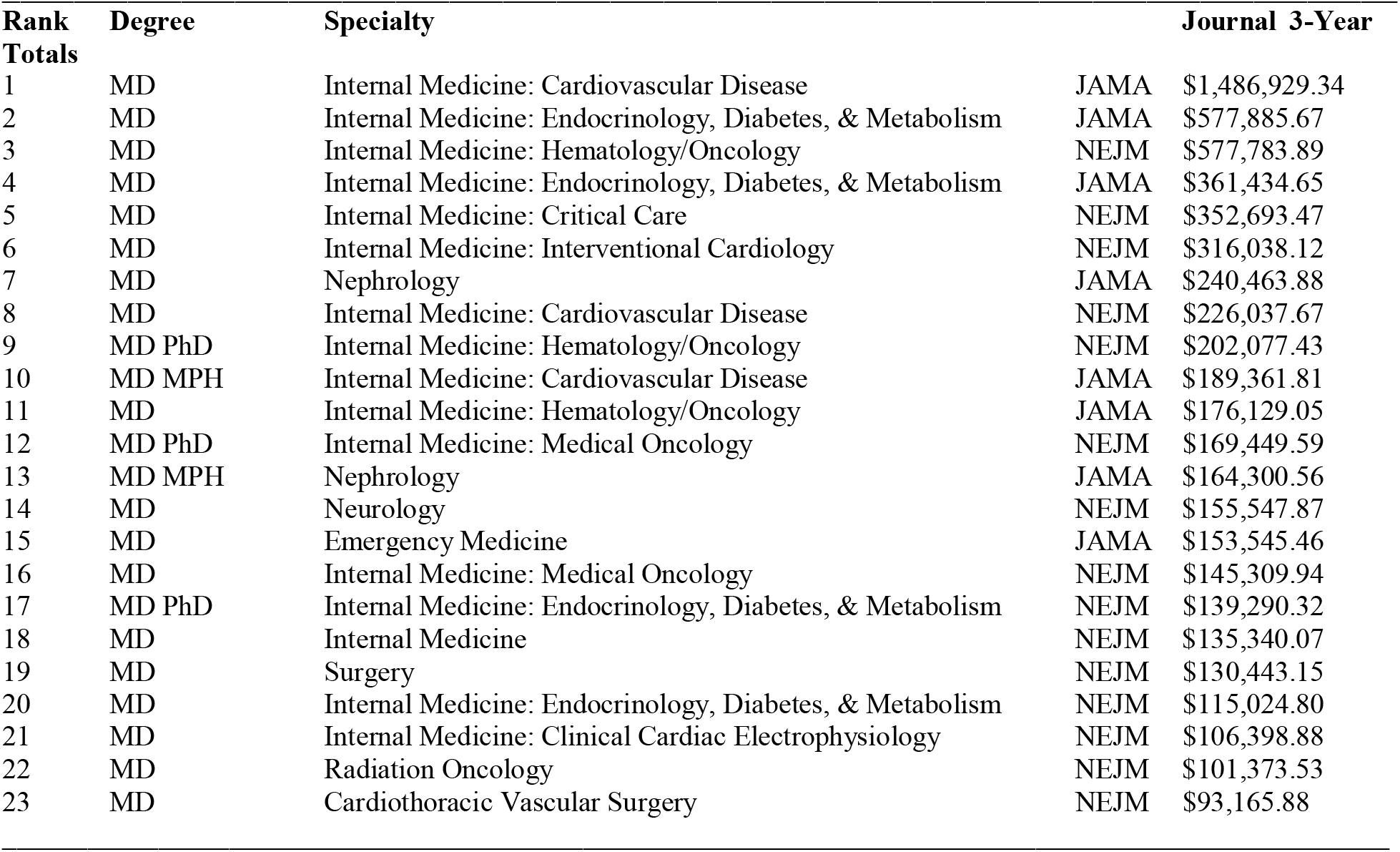
Characteristics of the top 23 highest earning doctors (statistical outliers). 3-Year Total refers to the total amount within the 36-month disclosure window.

### COI Disclosure Rates

Of the 106 authors who received payments, 55 made disclosures of which the disclosed companies reported dollar amounts that summed to at least half of the authors’ total payment amounts. Twenty had a three-year disclosure rate of 100%; ten of these published in *NEJM*, and the other ten published in *JAMA*. The other 35 authors who disclosed at least half of their payments had disclosure rates that ranged from 54.5% to 99.9%. Of the 51 authors who disclosed less than half of their payment amounts, eighteen disclosed between 0.007% and 42.3%. 33 authors who received payments disclosed 0%, or no amount, of their payments received. Of the authors who disclosed 0%, 21 of them published in *JAMA*, and twelve published in *NEJM* (Figure 3).

**FIGURE 3.**
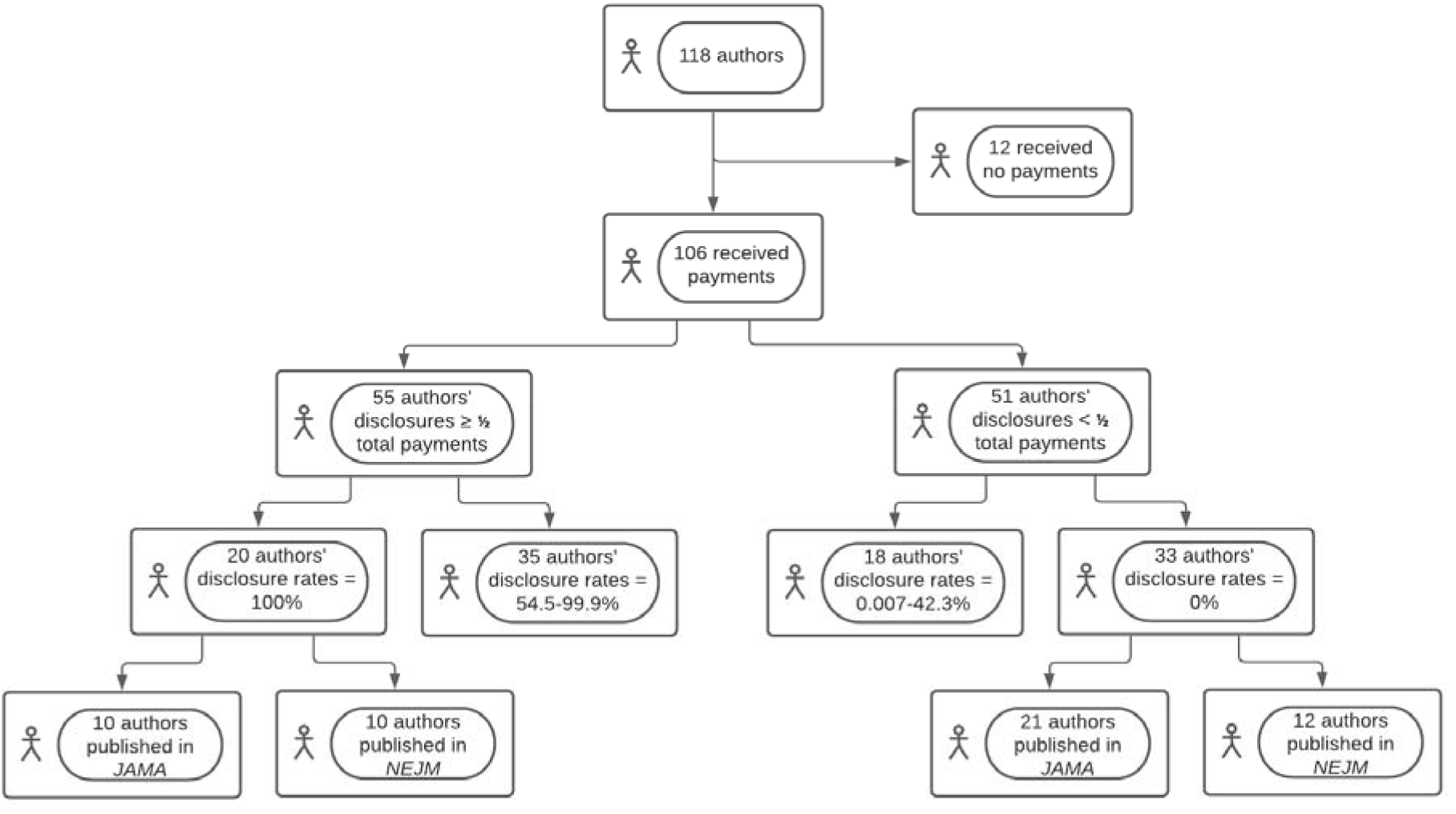
Flowchart of payment disclosure rate distributions. Of the 35 authors who disclosed at least half (but not 100%) of their payment amounts, the range of actual disclosure rates was 54.5 to 99.9%. Of the eighteen authors who disclosed less than half (but not 0%) of their payment amounts, the range of actual disclosure rates was 0.007 to 42.3%.

Subgroup analysis further examined the disclosure rates of all physician-authors who received compensation by comparing the outliers with the non-outliers. A Wilcoxon rank-sum test found no significant difference (*p* = 0.1849) between these two subgroups. The median disclosure rate of outliers was 78.1% and of non-outliers was 33.4%.

Sensitivity analysis compared the disclosure rates of all physician-authors who received compensation with those compensated who were not outliers. A Wilcoxon rank-sum test found no significant difference (*p* = 0.6406) between these two groups. The median disclosure rate of all those who received compensation was 68.1%, while the median disclosure rate of non-outliers was 33.4%.

A Wilcoxon rank-sum test also found no significant difference in COI disclosure rates between *NEJM* and *JAMA* authors (*p* = 0.0849). The median disclosure rate of *NEJM* authors was 79.4% and of *JAMA* authors was 3.50%.

### COI by Year

Little variability was observed across the individual years that fell within the 36-month disclosure window. In 2014, 79 authors (67.0%) received payments; in 2015, 84 (71.2%) received payments; and in 2016, 81 (68.6%) received payments. In all three years the majority of physician-authors received no payments in the disclosed, indeterminate, and unrelated categories, whereas the majority did receive undisclosed payments in each year. Some authors who received no payments in one year within the disclosure window received payments in one or more of the other two years (Supplementary Materials).

### COI by Specialty

The physician-authors in this study represented 33 distinct medical specialties. Fourteen (42.4%) of these specialties were subspecialties of internal medicine. The two most-represented specialties among the 118 physician-authors were cardiovascular disease (*n* = 16) and general internal medicine (*n* = 11). Ten specialties each were represented by four to nine individuals, seven specialties each were represented by three individuals, five specialties each by two individuals, and eleven specialties each by a single individual (Supplementary Materials).

## DISCUSSION

These novel data from highly influential US general medical journals (*NEJM* and *JAMA*) extend a sizable evidence base that has raised doubts about whether self-reported financial disclosure is a trustworthy mechanism for point-of-care databases,^13^ clinical practice guidelines,^33,34^ or other authoritative resources.^3^ Financial COIs are important to identify in order to recognize sources of potential bias in research works published by physicians and other researchers. Such bias can have devastating consequences: it undermines public trust in science,^35-38^ confounds understanding of treatment efficacy^39-41^ and clinical practice guidelines,^9-13^ and even continues to obstruct investigation into the origins of SARS-CoV2.^42,43^ Such instances provoke popular outrage^44-46^ and incite corrective action,^47,48^ often to little avail.^29,49^ The May 2, 2017, *JAMA* “theme” issue that dedicated a variety of articles to the topic of COI disclosures.^49^ On multiple occasions, both *NEJM* and *JAMA*, as well as many other publications, have confronted the resignation or dismissal of their editors-in-chief for COIs, financial and otherwise.^50-54^ Perhaps *JAMA*’s submitting authors have exercised greater COI disclosure transparency since publication of that 2017 special issue, but the results of the present study are not consistent with this supposition.

COI disclosure rules and procedures venture to mitigate the impact of COI bias on the integrity of published manuscripts. This premise means that ascertaining the impact of payments on researchers, or how payments influence those receiving them, may help to delineate the process of this insult to publication integrity. Key to identifying such bias in the first place is examination of COI disclosure accuracy. This was the purpose of the present study. Accordingly, the authors of this study take no position on the physician-authors’ intentions in non-disclosure of their COIs; we characterize the issue as a “process problem” rather than a “people problem,” especially in light of the patterns observed in COI disclosure rates regardless of the journal’s disclosure process and regardless of payment amount.

It was hypothesized that different disclosure processes between the two journals could produce different patterns in payment distributions and disclosure rates. This hypothesis derives from the fact that *NEJM* provided a copy of the original disclosure, while *JAMA* provided a list of disclosures. On the contrary, the data demonstrate no such significant differences between the two journals. Both journals had higher payment concentrations amongst the outliers than amongst the rest of the sample, illustrating a Pareto-principle pattern reflected across the two journals. This lack of differences in payment distributions and disclosure rates, despite a difference in the disclosure process, may imply that the journals’ differing disclosure processes had no effect on payment distributions and disclosure rates.

Further analysis elucidated surprising parity between the outliers and the entire study sample. Subgroup analysis indicated that physician-authors were just as likely to disclose COIs if their payment amounts did or did not constitute an outlier, and sensitivity analysis indicated that non-outliers were just as likely to disclose COIs as the entire group of physician-authors who received compensation. Therefore, COI disclosure rates remained consistent regardless of total payment amount, answering the question posed by previous researchers of the difference that payment amount makes in COI disclosure.^55^ This suggests that adherence to, or non-compliance with, COI disclosure procedures was not a function of payment amount, even for the outliers. It follows that targeting for correction the COI disclosure practices of those receiving the highest payment amounts would not address the fundamental problem of COI non-disclosure that this study finds to be common to physician-authors who have received general-payment compensation.

A future study comparing COI disclosure patterns with this earliest period of Open Payments data might show a change in such patterns and, possibly, the effect of Open Payments on COI disclosure transparency. Future research may be more robust now that the SUPPORT Act has expanded the range of researchers whose data are collected by Open Payments.^56^ As of January, 2021, physician assistants, clinical nurse specialists, certified nurse midwives, certified registered nurse anesthetists, and anesthesiologist assistants have entries on the Open Payments website.^57^ This new data source may help to assess whether the observations of this study are applicable to mid-level healthcare providers.

### Limitations

The major limitation of this study is that of generalizability, especially of the findings to journals other than *NEJM* and *JAMA*. The authors assessed the data within a non-parametric analytical framework because there is no methodological justification for making inferences about payment distribution patterns amongst the broader community of physician-authors. The 118 physician-authors that met inclusion criteria are not presumed to represent this broader community, despite the more general-interest nature of the content that *NEJM* and *JAMA* tend to publish. Moreover, the authors who publish in these two journals may more likely receive funding than those who publish in other journals generally. This suggests that the data may be skewed towards those who already receive more compensation, although this study’s findings of no difference in disclosure rates between outliers and non-outliers may belie this point. Another limitation was small sample size across the two journals, which may have obscured possible differences due to low statistical power.

## CONCLUSION

The fact that the preponderance (81.1%) of physician-authors in this study received payments that they did not disclose but that they nonetheless were supposed to disclose as COIs per ICMJE guidelines and journal requirements demonstrates that these disclosure requirements in conjunction with the expectation of COI self-disclosure have been inadequate to ensure full COI transparency in either *NEJM* or *JAMA* and regardless of general payment amount received. Making industry payments a matter of public record in the form of Open Payments presumes to mitigate this problem of COI disclosure opacity. Readers are encouraged to “trust, but verify” and to compare self-reported with industry-reported disclosures early in the investigative process.

## Supporting information

Raw data

## Data Availability

Data is included as a supplement.

## DISCLOSURES

### Sources of Support (if applicable)

This research received no specific grant from any funding agency in the public, commercial, or not-for-profit sectors.

## SUPPLEMENTARY MATERIALS

See Spreadsheet for full data.

See ICMJE Form for Disclosure of Potential Conflicts of Interest.

## REFERENCES

1. Torgerson T, Wayant C, Cosgrove L, Akl EA, Checketts J, Dal Re R, Gill J, Grover SC, Khan N, Khan R, Marušić A, McCoy MS, Mitchell A, Prasad V, Vassar M. Ten years later: a review of the US 2009 Institute of Medicine report on conflicts of interest and solutions for further reform. BMJ Evid Based Med. 2020;bmjebm-2020–111503.

2. Bekelman JE, Li Y, Gross CP. Scope and impact of financial conflicts of interest in biomedical research: a systematic review. JAMA. 2003;289(4):454–465.

3. Krimsky S, Schwab T. Conflicts of interest among committee members in the National Academies’ genetically engineered crop study. PLoS ONE. 2017;12(2):e0172317.

4. Lundh A, Lexchin J, Mintzes B, Schroll JB, Bero L. Industry sponsorship and research outcome. Cochrane Database Syst Rev. 2017;2:MR000033.

5. Yeh JS, Franklin JM, Avorn J, Landon J, Kesselheim AS. Association of industry payments to physicians with the prescribing of brand-name statins in Massachusetts. JAMA Intern Med. 2016;176(6):763.

6. Hadland SE, Rivera-Aguirre A, Marshall BD, Cerdá M. Association of pharmaceutical industry marketing of opioid products with mortality from opioid-related overdoses. JAMA Netw. 2019;2(1):e186007.

7. Nusrat S, Syed T, Nusrat S, Chen S, Chen W-J, Bielefeldt. Assessment of pharmaceutical company and device manufacturer payments to gastroenterologists and their participation in clinical practice guideline panels. JAMA Netw. 2018;1(8):e186343.

8. Tanne JH. US makers of joint replacements are fined for paying surgeons to use their devices. BMJ. 2007;335(7629):1065.

9. Cosgrove L, Bursztajn HJ, Krimsky S, Anaya M, Walker J. Conflicts of interest and disclosure in the American Psychiatric Association’s Clinical Practice Guidelines. Psychother Psychosom. 2009;78(4):228–232.

10. Neuman J, Korenstein D, Ross JS, Keyhani S. Prevalence of financial conflicts of interest among panel members producing clinical practice guidelines in Canada and United States: cross sectional study. BMJ. 2011;343:d5621.

11. Norris SL, Holmer HK, Ogden LA, Burda BU. Conflict of interest in clinical practice guideline development: a systematic review. PLoS One. 2011;6(10):e25153.

12. Piper BJ, Alinea AA, Wroblewski JR, Graham SM, Chung DY, McCutcheon LRM, Birkett MA, Kheloussi SS, Shah VM, Szarek JL, Zalim QK, Arnott JA, McLaughlin WA, Lucchesi PA, Miller KA, Waite GN, Bordonaro M. A quantitative and narrative evaluation of Goodman and Gilman’s Pharmacological Basis of Therapeutics. Pharmacy. 2019;8(1):1–20.

13. Chopra AC, Tilberry SS, Sternat KE, Chung DY, Nichols SD, Piper BJ. Quantification of conflicts of interest in an online point-of care clinical support website. Sci Eng Ethics. 2020;26(2):921–930.

14. Tabatabavakili S, Khan R, Scaffidi MA, Gimpaya N, Lightfoot D, Grover SC. Financial conflicts of interest in clinical practice guidelines: a systematic review. Mayo Clin Proc Innov Qual Outcomes. 2021;5(2):466–475.

15. Traversy G, Barnieh L, Akl EA, Allan GM, Brouwers M, Ganache I, Grundy Q, Guyatt GH, Kelsall D, Leng G, Moore A, Persaud N, Schünemann HJ, Straus S, Thombs BD, Rodin R, Tonelli M. Gestion des conflits d’intérêts durant l’élaboration de lignes directrices en santé [Managing conflicts of interest during the development of health guidelines]. CMAJ. 2021;193(9):E324–E330.

16. Brems JH, Davis AE, Clayton EW. Analysis of conflict of interest policies among organizations producing clinical practice guidelines. PLoS One. 2021;16(4):e0249267.

17. Congress.gov. S.301 - Physician Payments Sunshine Act of 2009. https://www.congress.gov/111/bills/s301/BILLS-111s301is.pdf. Accessed September 5, 2021.

18. Centers for Medicare and Medicaid Services. What is Open Payments? https://www.cms.gov/OpenPayments. Accessed September 5, 2021.

19. ICMJE. Journals stating that they follow the ICMJE Recommendations. http://www.icmje.org/journals-following-the-icmje-recommendations/. Accessed September 5, 2021.

20. Checketts JX, Sims MT, Vassar M. Evaluating industry payments among dermatology clinical practice guidelines authors. JAMA Dermatol. 2017;153(12):1229–1235.

21. Horn J, Checketts JX, Jawhar O, Vassar M. Evaluation of industry relationships among authors of otolaryngology clinical practice guidelines. JAMA Otolaryngol Head Neck Surg. 2018;144(3):194–201.

22. Ziai K, Pigazzi A, Smith BR, Nouri-Nikbakht R, Nepomuceno H, Carmichael JC, Mills S, Stamos MJ, Jafari MD. Association of compensation from the surgical and medical device industry to physicians and self-declared conflict of interest. JAMA Surg. 2018;153(11):997–1002.

23. Waqas A, Baig AA, Khalid MA, Aedma KK, Naveed S. Conflicts of interest and outcomes of clinical trials of antidepressants: an 18-year retrospective study. J Psychiatr Res. 2019;116:83–87.

24. Wayant C, Turner E, Meyer C, Sinnett P, Vassar M. Financial conflicts of interest among oncologist authors of reports of clinical drug trials. JAMA Oncol. 2018;4(10):1426–1428.

25. Ozaki A. Conflict of interest and the CREATE-X Trial in the New England Journal of Medicine. Sci Eng Ethics. 2018;24(6):1809–1811.

26. Goldner JA. Dealing with conflicts of interest in biomedical research: IRB oversight as the next best solution to the abolitionist approach. J Law Med Ethics. 2000;28(4):379–404.

27. Novins DK, Althoff RR, Billingsley MK, Cortese S, Drury SS, Frazier JA, Henderson SW, McCauley EA, White TJH, Karnik NS. Conflict of interest and the Journal revisited. J Am Acad Child Adolesc Psychiatry. 2018;57(2):72–73.

28. Cherla DV, Olavarria OA, Holihan JL, Viso CP, Hannon C, Kao LS, Ko TC, Liang MK. Discordance of conflict of interest self-disclosure and the Centers of Medicare and Medicaid Services. J Surg Res. 2017;218:18–22.

29. Bauchner H, Fontanarosa PB, Flanagin A. Conflicts of interests, authors, and journals: new challenges for a persistent problem. JAMA. 2018;320(22):2315–2318.

30. U.S. Centers for Medicare & Medicaid Services. Search Open Payments. https://openpaymentsdata.cms.gov/. Accessed September 5, 2021.

31. ICMJE. Disclosure of Interest (Updated February 2021). http://www.icmje.org/disclosure-of-interest/. Accessed September 5, 2021.

32. JAMA. For Authors. https://jamanetwork.com/journals/jama/pages/for-authors. Accessed September 5, 2021.

33. Murayama A, Ozaki A, Saito H, Sawano T, Shimada Y, Yamamoto K, Suzuki Y, Tanimoto T. Pharmaceutical company payments to dermatology clinical practice guideline authors in Japan. PLoS One. 2021;15(10):e0239610.

34. Bansal R, Khan R, Scaffidi MA, Gimpaya N, Genis S, Bukhari A, Dhillon J, Dao K, Bonneau C, Grover SC. Undisclosed payments by pharmaceutical and medical device manufacturers to authors of endoscopy guidelines in the United States. Gastrointest Endosc. 2020;91(2):266–273.

35. Davidoff F, DeAngelis CD, Drazen JM, Hoey J, Højgaard L, Horton R, Kotzin S, Nicholls MG, Nylenna M, Overbeke AJPM, Sox HC, Van Der Weyden MB. Sponsorship, authorship, and accountability. NEJM. 2001;345:825–827.

36. Smith R. Research misconduct: the poisoning of the wells. J Royal Soc Med. 2006;99:237f.

37. Snyder PJ, Mayes LC, Spencer D. Science and the Media: Delgado’s Brave Bulls and the Ethics of Scientific Disclosure. Academic Press: London, UK. 2009.

38. Cigarroa FG, Masters BS, Sharphorn D. Institutional conflicts of interest and public trust. JAMA. 2018;320(22):2305–2306.

39. Armstrong D. Delicate operation: how a famed hospital invests in a device it uses and promotes. Wall Street Journal. A1. December 12, 2005.

40. Armstrong D. Drug interactions: financial ties to industry cloud major depression study. Wall Street Journal. A1. July 11, 2006.

41. Lo B, Field MJ (eds.). Conflict of Interest in Medical Research, Education, and Practice. National Academic Press: Washington, DC. 2009.

42. Editors of The Lancet. Addendum: competing interests and the origins of SARS-CoV-2. Lancet. 2021;397(10293):2449–2450.

43. Editorial Board. Who are the COVID investigators? Wall Street Journal. February 15, 2021.

44. Matthews D. Under-fire Lancet admits conflict of interest on lab-leak letter. Times Higher Education. June 22, 2021.

45. Geraghty J. China apologist Peter Daszak has some explaining to do. National Review. June 22, 2021.

46. Spence M. The rise and fall of British virus hunter Peter Daszak. The Times. June 26, 2021.

47. Guyatt G, Akl EA, Hirsh J, Kearon C, Crowther M, Gutterman Lewis SZ, Nathanson I, Jaeschke R, Schünemann H. The vexing problem of guidelines and conflict of interest: a potential solution. Ann Intern Med. 2010;152(11):738–741.

48. Cosgrove L, Bursztajn HJ, Erlich DR, Wheeler EE, Shaughnessy AF. Conflicts of interest and the quality of recommendations in clinical guidelines. J Eval Clin Pract. 2013;19(4):674–681.

49. Fontanarosa P & Bauchner H. Conflict of interest and medical journals. JAMA. 2017;317(17):1768–1771.

50. Mississippi Valley Medical Association. A medical editor’s resignation. JAMA. 1893;21(16):582.

51. Brown D. JAMA Editor fired over Clinton conflict. Washington Post. A03. January 16, 1999.

52. Stalman WA. 1999, het jaar van de ontslagen hoofdredacteuren [1999, the year of fired editors-in-chief]. Ned Tijdschr Geneeskd. 2000;144(9):447–448.

53. Pincock S. Journal editor quits in conflict scandal. The Scientist. August 27, 2006. https://www.the-scientist.com/daily-news/journal-editor-quits-in-conflict-scandal-47277. Accessed September 5, 2021.

54. Ferguson C, Marcus A, Oransky I. Publishing: The peer-review scam. Nature. 2014;515:480–482.

55. Lo B, Grady D. Payments to physicians: does the amount of money make a difference? JAMA. 2017;317(17):1719–1720.

56. Congress.gov. Public Law 115-271 – Oct. 24, 2018 – Substance Use-Disorder Prevention That Promotes Opioid Recovery and Treatment for Patients and Communities Act (SUPPORT for Patients and Communities Act). https://www.congress.gov/115/plaws/publ271/PLAW-115publ271.pdf. Accessed September 5, 2021.

57. Open Payments Law and Policy. CMS. https://www.cms.gov/OpenPayments/Law-and-Policy. Accessed September 5, 2021.

